# Prevalence of Venous Thromboembolism in Critically-ill COVID-19 Patients: Systematic Review and Meta-analysis

**DOI:** 10.1101/2020.08.24.20175745

**Authors:** Mouhand F.H. Mohamed, Shaikha D. Al-Shokri, Khaled M. Shunnar, Sara F. Mohamed, Mostafa S. Najim, Shahd I. Ibrahim, Hazem Elewa, Khalid M. Dousa, Lina O. Abdalla, Ahmed El-Bardissy, Mohamed Nabil Elshafei, Ibrahim Y. Abubeker, Mohammed Danjuma, Mohamed A Yassin

## Abstract

**Background:** Recent studies revealed a high prevalence of venous thromboembolism (VTE) events in coronavirus disease 2019 (COVID-19) patients, especially in those who are critically ill. Available studies report varying prevalence rates. Hence, the exact prevalence remains uncertain. Moreover, there is an ongoing debate regarding the appropriate dosage of thromboprophylaxis. Methods: We performed a systematic review and proportion meta-analysis following PRISMA guidelines. We searched PubMed and EMBASE for studies exploring the prevalence of VTE in critically ill COVID-19 patients till 22/07/2020. We pooled the proportion of VTE. Additionally, in a subgroup analysis, we pooled VTE events detected by systematic screening. Finally, we compared the odds of VTE in patients on prophylactic compared to therapeutic anticoagulation. Results: The review comprised of 24 studies and over 2500 patients. The pooled proportion of VTE prevalence was 0.31 (95% CI 0.24, 0.39 I^2^ 94%), of VTE utilizing systematic screening was 0.48 (95% CI 0.33, 0.63 I^2^ 91%), of deep-venous-thrombosis was 0.23 (95% CI 0.14, 0.32 I^2^ 96%), of pulmonary embolism was 0.14 (95% CI 0.09, 0.20 I^2^ 90%). In a subgroup of studies, utilizing systematic screening, VTE risk increased significantly with prophylactic, compared to therapeutic anticoagulation (OR 5.45; 95% CI 1.90, 15.57 I^2^ 0%). Discussion: Our review revealed a high prevalence of VTE in critically ill COVID-19 patients. Almost 50% of patients had VTE detected by systematic screening. Higher thromboprophylaxis dosages seem to reduce VTE burden in this patient’s cohort compared to standard prophylactic anticoagulation; ongoing randomized controlled trials will further confirm this.

## Introduction

The pool of recent evidence suggests that Coronavirus disease 2019 (COVID-19) is a thrombogenic condition. It leads to an increased incidence of both venous and arterial thromboembolic events.^1^ COVID-19 patients admitted to intensive care units (ICU) seem to carry a higher risk.^1^ Venous thromboembolism (VTE) prevalence in the critically ill COVID-19 patients varied across individual studies. This is likely due to differences in screening methods (systematic vs. nonsystematic screening), among other study-specific characteristics, leaving VTE’s exact prevalence unknown. The prevalence of deep venous thrombosis (DVT) was considered low compared to pulmonary embolism (PE), which led researchers to consider micro thrombosis as an additional mechanism of PE in COVID-19 patients.^2^

VTE’s heightened risk led to a wide chemoprophylaxis use for critically ill COVID-19 patients ^3^. Notwithstanding this, recent studies showed that even COVID-19 patients on chemoprophylaxis remain to carry a high risk of VTE compared to non-COVID-19 patients.^4^ As a result, guidance driven by expert opinions suggested utilizing higher doses of anticoagulation.^1^ However, this recommendation lacks robust, supporting systematic studies. Thus, we aimed to systematically review the literature and explore the pooled prevalence of VTE, PE, and DVT in critically ill COVID-19 patients. Additionally, to evaluate the yield of systematic VTE screening and its effect on the prevalence. Moreover, if data allow, to examine the odds of VTE in patients on prophylactic compared to therapeutic anticoagulation.

## Methods

This review follows the Preferred Reporting Items for Systematic Reviews and Meta-Analyses (PRISMA) guidelines.^5^ It is pre-registered at the International Prospective Register of Systematic Reviews (PROSPERO) (registration number: CRD42020185916).

### Eligibility criteria

We limited our review to observational studies (cohort, cross-sectional, retrospective, or case series) estimating the proportion of VTE events in critically ill COVID-19 adult (>18 years) patients (admitted to ICU). To facilitate a timely review, we limited our inclusion to articles written in the English language only. We excluded studies where the proportion of VTE could not be ascertained, or if the population of interest is not ICU patients.

### Information sources and literature search

For a timely review, we performed the search in PubMed, MEDLINE, and EMBASE. We used free text, emtree, and MeSH terms in our search. There was no language or date limitations implied in the search; The last date of the formal search was the 10^th^ of July 2020; however, we performed a scoping search till July 25, 2020. Example of a utilized search strategy was ((‘venous thromboembolism’ OR ‘deep vein thrombosis’ OR ‘lung embolism’ OR ‘vein thrombosis’/exp/mj) AND [embase]/lim) AND ((‘covid 19’ OR (coronavirus AND disease AND 2019) OR (sars AND cov AND 2) OR ‘covid 19’/exp/mj) AND [embase]/lim). We also performed relevant citations and reference searches.

### Screening and data extraction

Two reviewers (MFHM) and (SFM) conducted the screening in two stages. The first stage was screening the retrieved articles’ titles and abstracts independently. Secondly, the articles’ full text was retrieved and assessed for inclusion. When disagreement occurred, a third reviewer (LOA) settled the disagreement guided by the protocol. We used pre-made excel sheets to collect relevant articles data. This included the last author name, publication date, study country, sample size, events number (DVT, PE, and VTE), baseline characteristics (median age, gender frequency, average BMI, and other comorbidities), intubation frequency, thromboprophylaxis frequency, and follow-up duration.

### Study quality and risk of bias assessment

We used a validated tool for assessing the risk of bias of prevalence studies. The tool was devised by Hoy et al. and is comprised of ten items summarizing four domains.^6^ We additionally generated funnel plots to examine the risk of publication bias in our review.

### Statistical analysis

A scoping review revealed heterogeneity of the method of VTE screening, reporting, and detection. Additionally, there were varying follow-ups given the nature of ICU admitted patients. Hence, neither the true incidence (different follow-up time and some patients may already have the event of interest before the study) nor the true prevalence (varying follow-up time and absence of unifying screening for all individuals at risk) could be accurately pooled. We instead decided a priori to pool a proportion of VTE with a 95% confidence interval. This proportion represents the number of patients with the event of interest divided by the study population at risk during the study regardless of their follow-up duration. We felt that this would be a proxy or an estimate of the prevalence. We used the validated method of double arcsine transformation to stabilize the variance and confine the confidence interval between 0-1.^7^ We generated forest plots to display the results of the analysis. We used the Cochrane Q test and I^2^ to examine heterogeneity. I^2^ >60% indicates significant heterogeneity. Regardless of the heterogeneity, we would use the random-effects model (REM) in our analysis. We used MetaXl software for statistical analysis (version 5.3 © EpiGear International Pty Ltd ABN 51 134 897 411 Sunrise Beach, Queensland, Australia, 2011-2016).

### Sub-group and sensitivity analyses

We a priori decided to examine the proportion of DVT and PE. Additionally, we looked at the proportion of VTE in various populations (systematic screening vs. nonsystematic screening, therapeutic vs. prophylactic anticoagulant dose). Moreover, we performed a sensitivity analysis to reflect the relative constituent studies’ impact on the consistency of the pooled proportion of the primary endpoint.

## Results

### Included studies and baseline characteristics

Twenty-four studies describing a total of 2570 patients were included in our final analysis (Figure 1 shows the flow diagram).^4,8–30^ The studies were heterogeneous in terms of VTE events identification and screening (Table 1). In ten studies, the screening for VTE was systematically done using lower and upper limb ultrasound (systematic screening was only for DVT and not PE). Fourteen studies evaluated for the presence of VTE based on clinical suspicion and further confirmation by imaging (nonsystematic). Twenty-two studies reported the proportion of DVTs, and seventeen studies reported the proportion of pulmonary embolism events. Out of the ten studies where systematic screening was adopted, the screening was incomplete in one. In all studies but one, ^26^ most patients were on thromboprophylaxis with varying doses.

**Figure 1:**
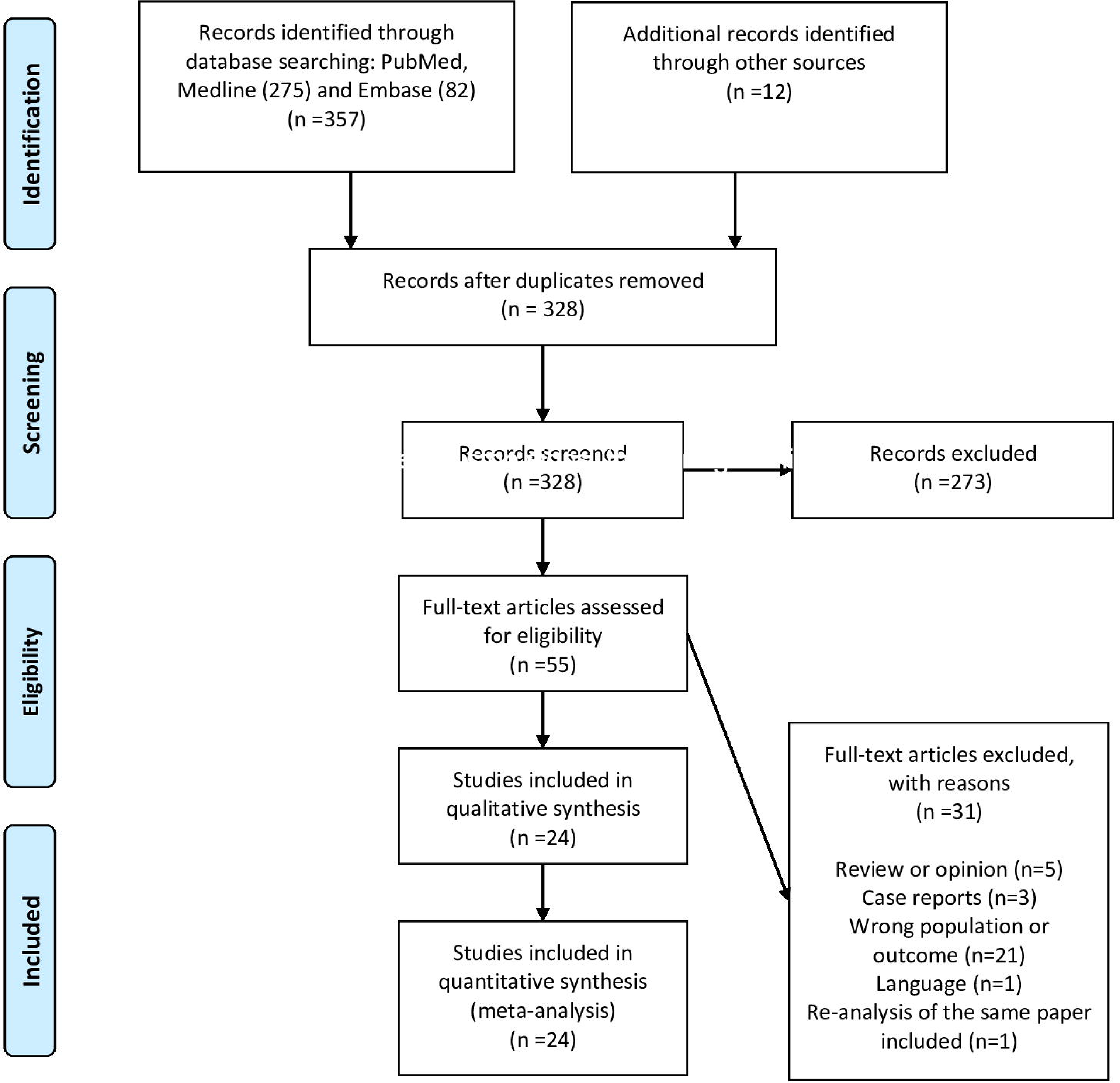
PRISMA flow diagram

**Table 1:**
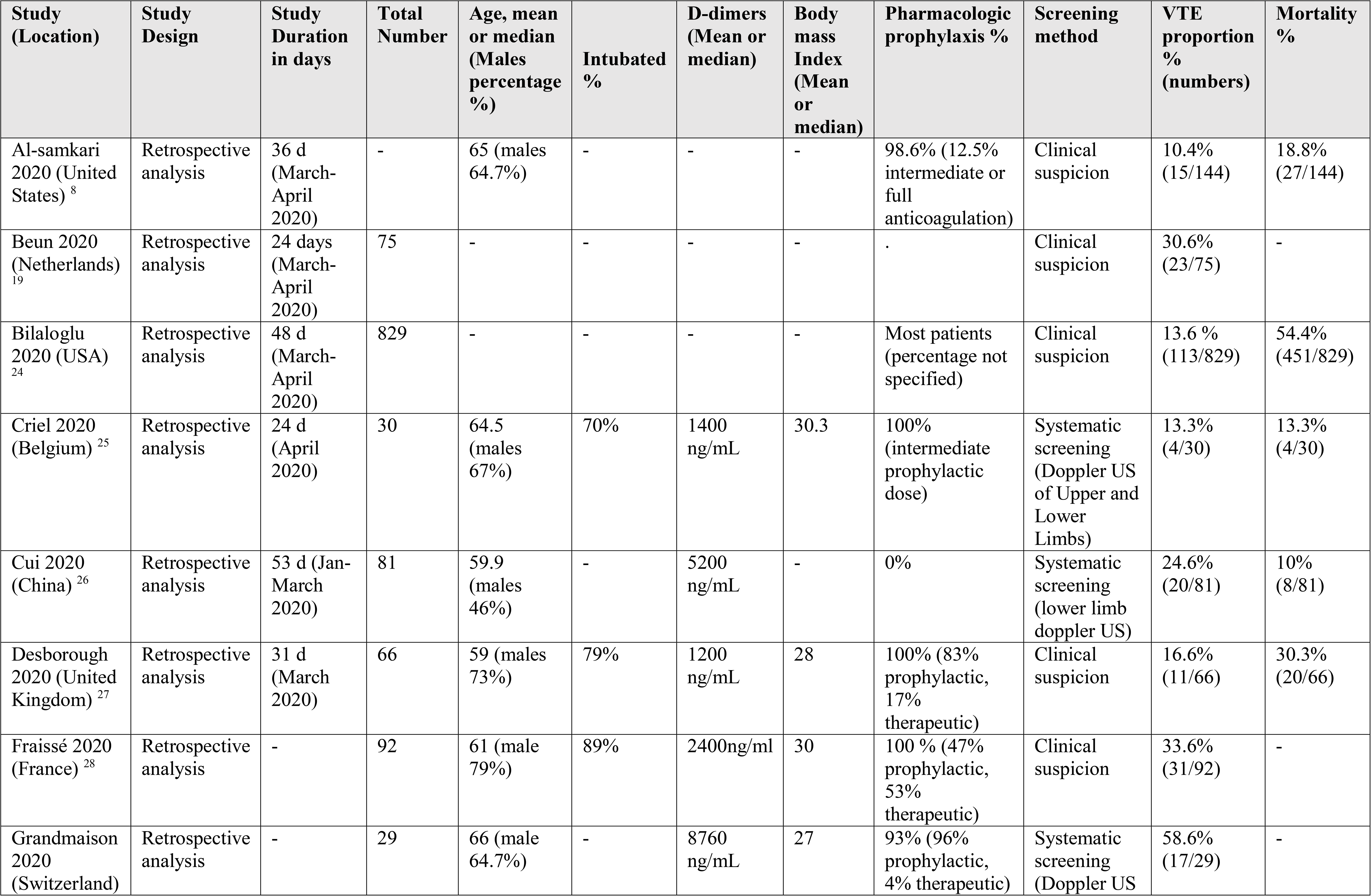
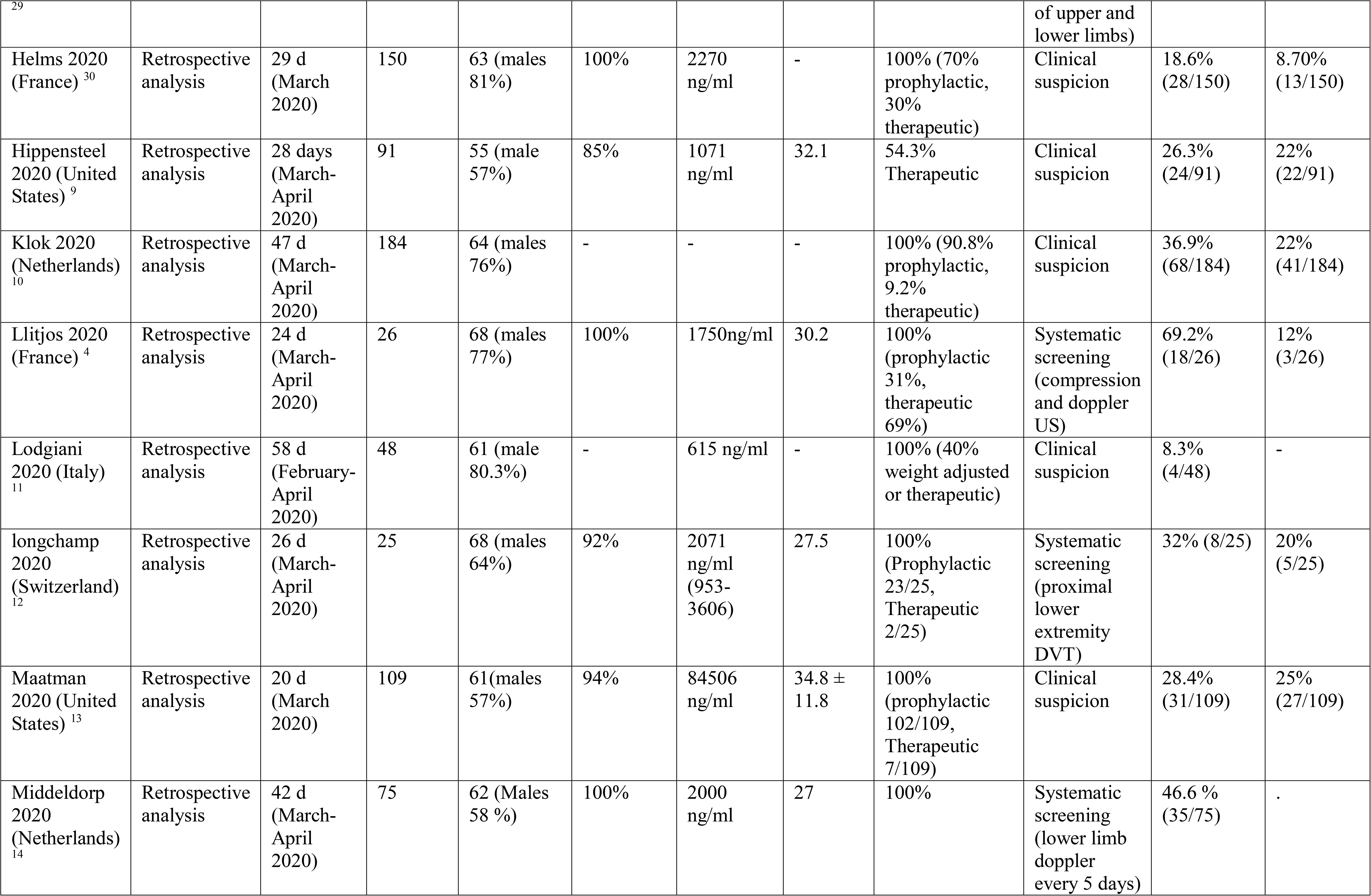
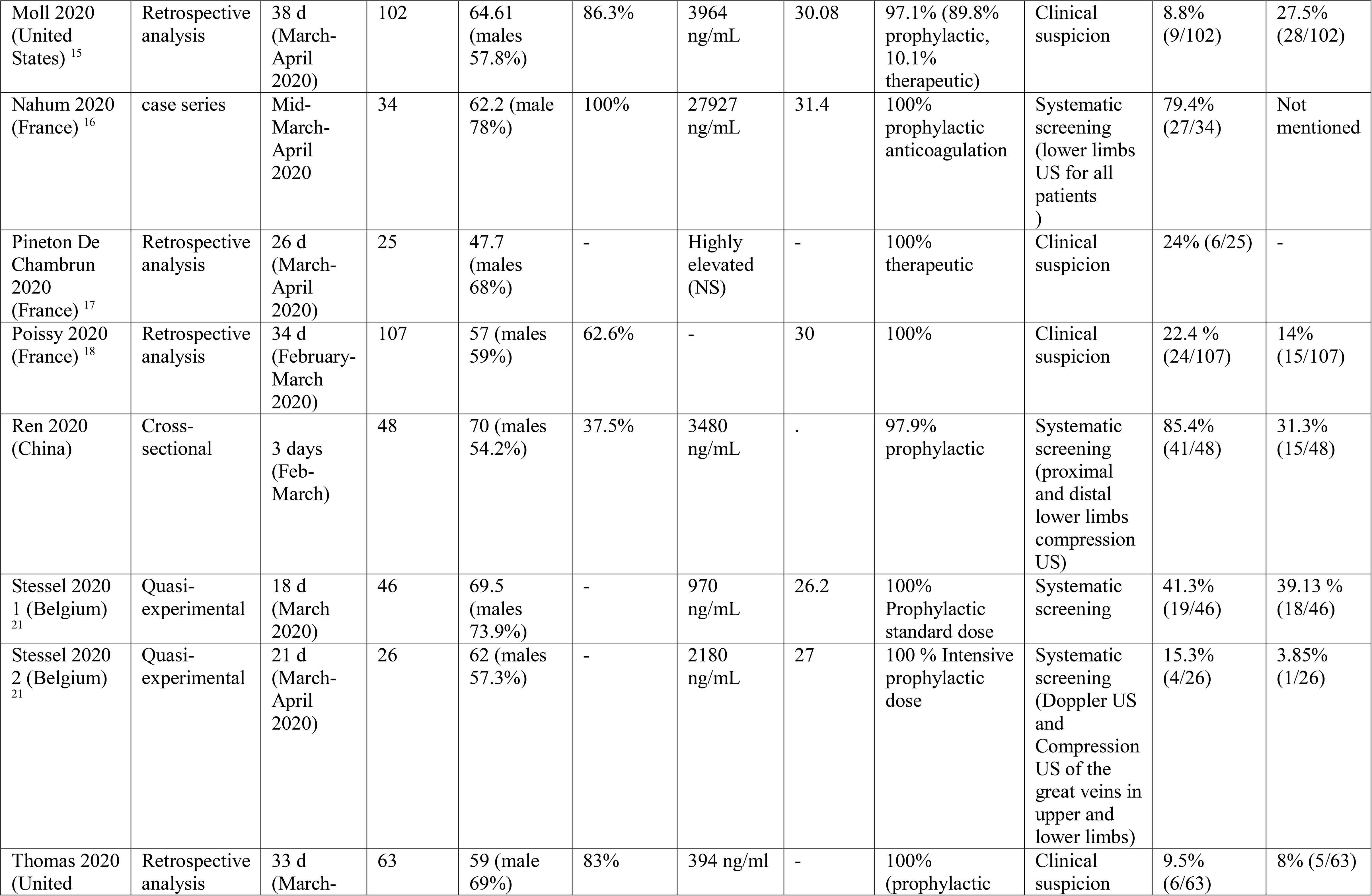
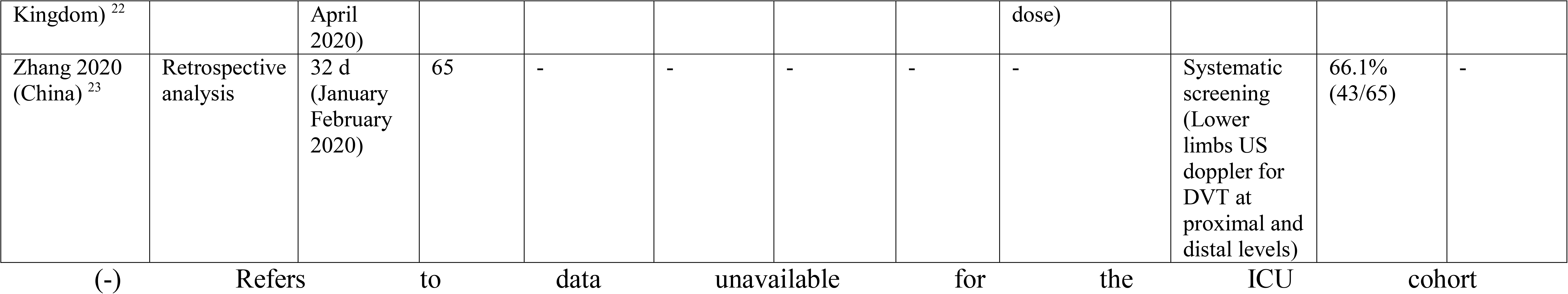
Summary of included studies

### The proportion of VTE events

The overall pooled proportion of VTE from 24 studies examining a total of 2570 was 0.31 (95% CI 0.24, 0.39 I^2^ 94% Q 383) with significant heterogeneity (Figure 2). The funnel plot showed significant asymmetry suggestive of possible publication bias (Supplementary S 1). The sensitivity analysis did not affect the final point estimate significantly (Supplementary S 2).

**Figure 2:**
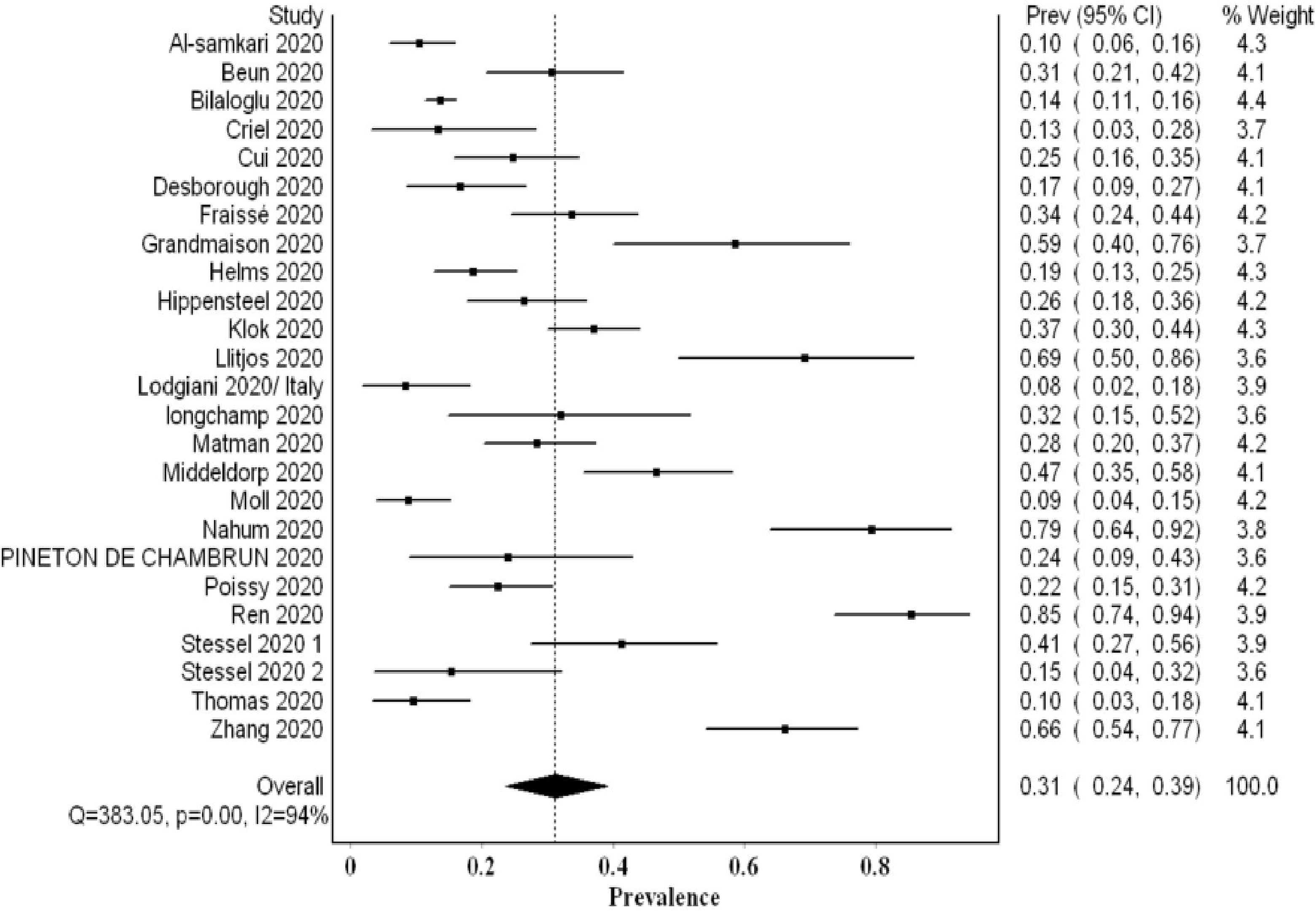
Forest plot showing the overall pooled proportion of VTE events

### The proportion of VTE utilizing systematic screening

Ten studies examining 478 patients using systematic screening, revealed a higher VTE proportion of 0.48 (95% CI 0.33, 0.63 I^2^ 91% Q 109) with significant heterogeneity (Figure 3). The funnel plot suggested a publication bias (Supplementary S3). The exclusion of Cui et al.’s study that did not utilize thromboprophylaxis resulted in a higher proportion of VTE events of 0.51. Additional sensitivity analyses revealed a lower VTE proportion with the exclusion of Ren et al.’s data (0.43); this proportion increased with the exclusion of Criel et al.’s study (0.52) (Supplementary S4). All the studies evaluated systematically for the presence of DVT events only (PE was not a primary aim). Hence, this pooled proportion represents the proportion of DVT events and may underestimate the overall VTE proportion.

**Figure 3:**
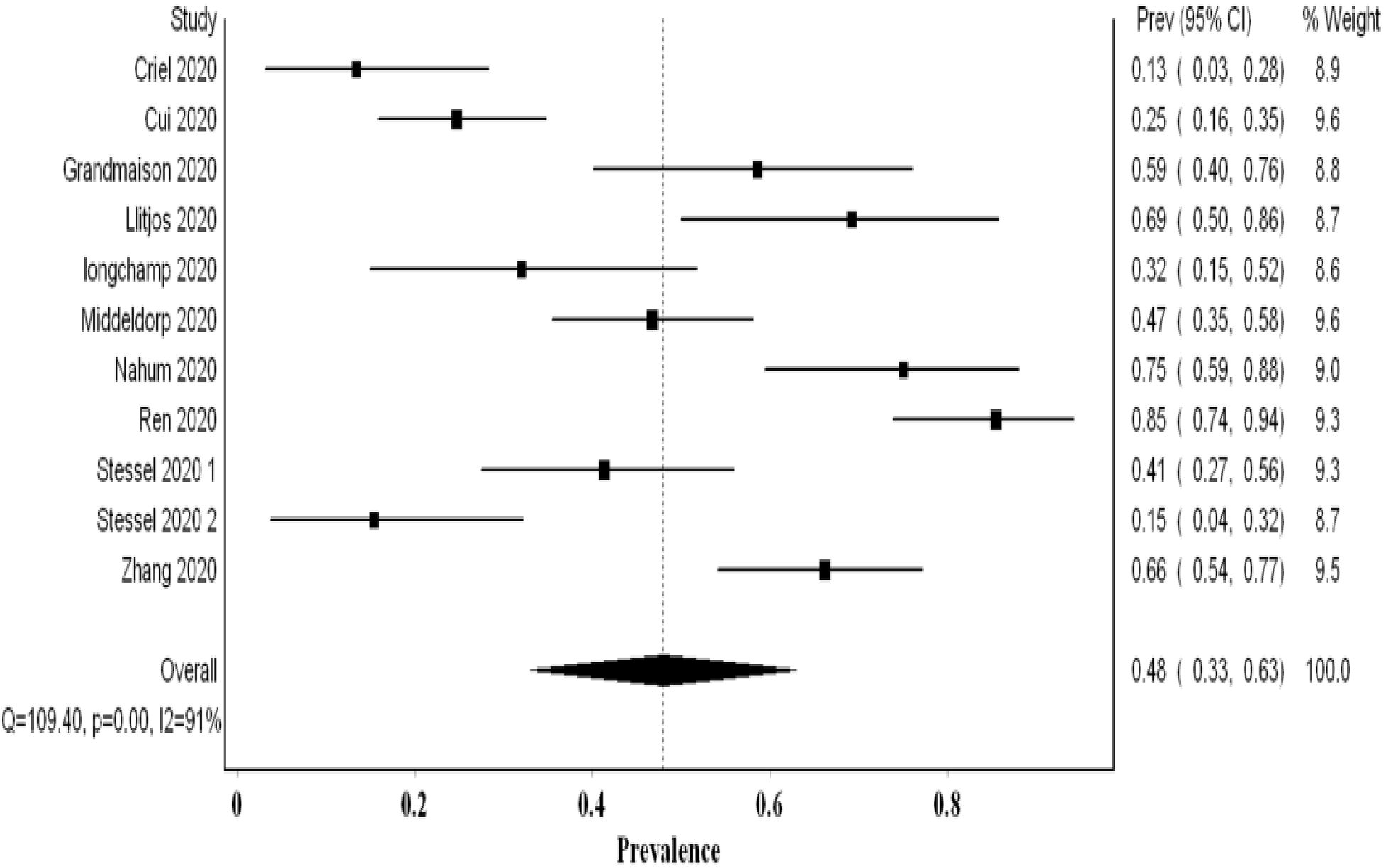
Forest plot showing the pooled proportion of VTE events utilizing systematic screening methods

### The proportion of VTE utilizing nonsystematic screening

In most studies utilizing nonsystematic screening, the authors addressed the high threshold for screening and imaging due to infection control implications. They stated that this might have underestimated the true prevalence. The analysis of fourteen studies examining 2085 patients revealed a pooled proportion of VTE of 0.20 (95% CI 0.15, 0.26 I^2^ 87% Q 98.4) (Figure 4). The funnel plot suggested a publication bias (Supplementary S 5). On sensitivity analysis, the final point estimate did not significantly change with the ordered exclusion of the constituent studies (Supplementary S 6).

**Figure 4:**
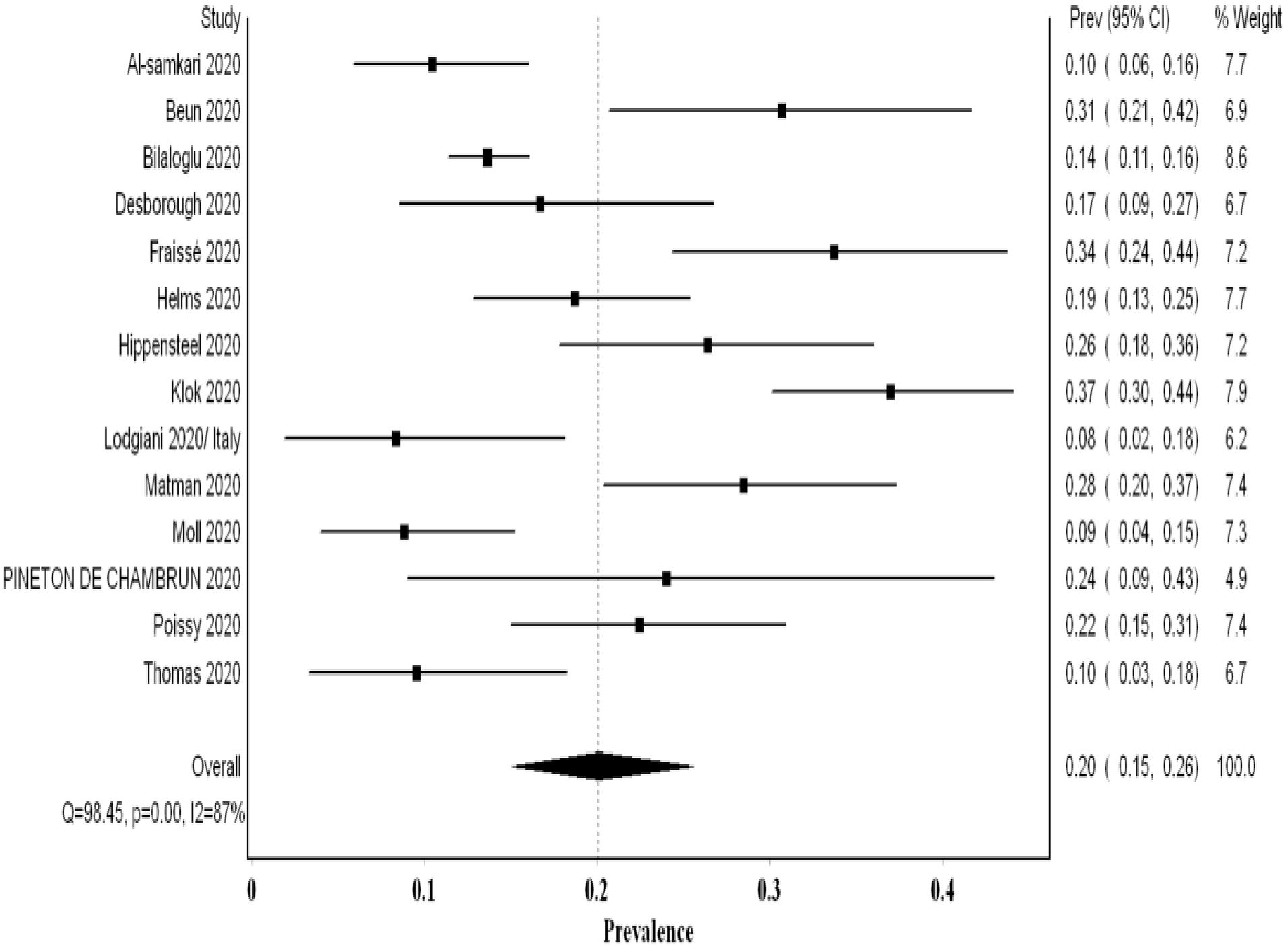
Forest plot showing the pooled proportion of VTE events utilizing non-systematic screening methods

### The proportion of DVT events

The overall pooled proportion of DVT from 22 studies examining a total of 2401 was 0.23 (95% CI 0.14, 0.32 I^2^ 96% Q 531) with significant heterogeneity (Figure 5). The funnel plot suggested a publication bias (S7). While the sensitivity analysis suggested a consistency of the final point estimate with ordered-single-study exclusion (Supplementary S8). The pooled proportion of DVT from studies utilizing nonsystematic screening was 0.08 (95% CI 0.04, 0.12 I^2^ 87% Q 85) (Supplementary S9).

**Figure 5:**
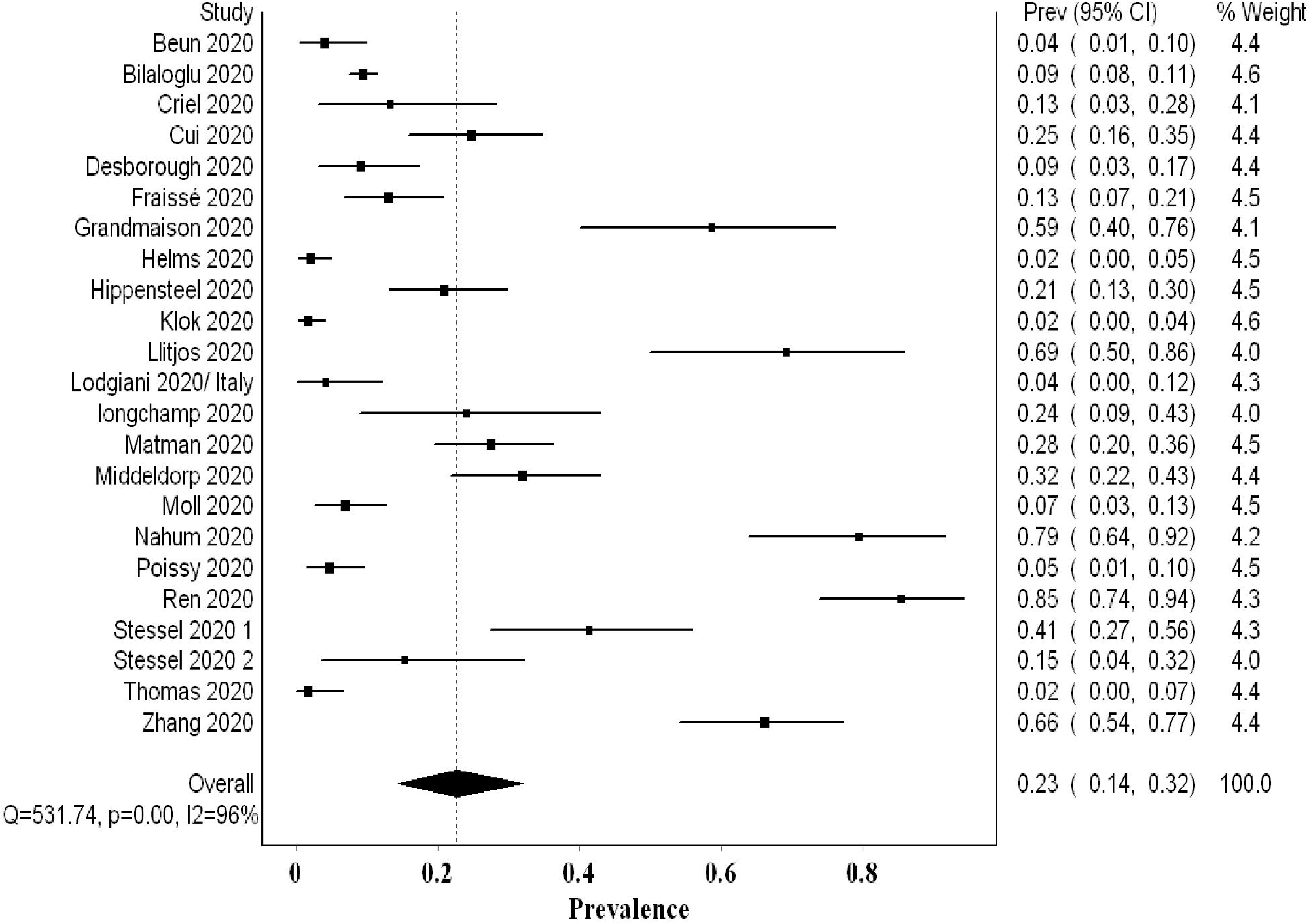
Forest plot showing the overall pooled proportion of DVT

### The proportion of PE events

Pulmonary embolism was not screened systematically. The analysis of 2096 patients (17 studies) revealed a pooled proportion of 0.14 (95% CI 0.09, 0.20 I^2^ 90% Q 159) (Figure 6). The funnel plot revealed a major asymmetry suggestive of publication bias (Supplementary S 10). Sensitivity analysis showed consistency of the results upon single-study-ordered exclusion.

**Figure 6:**
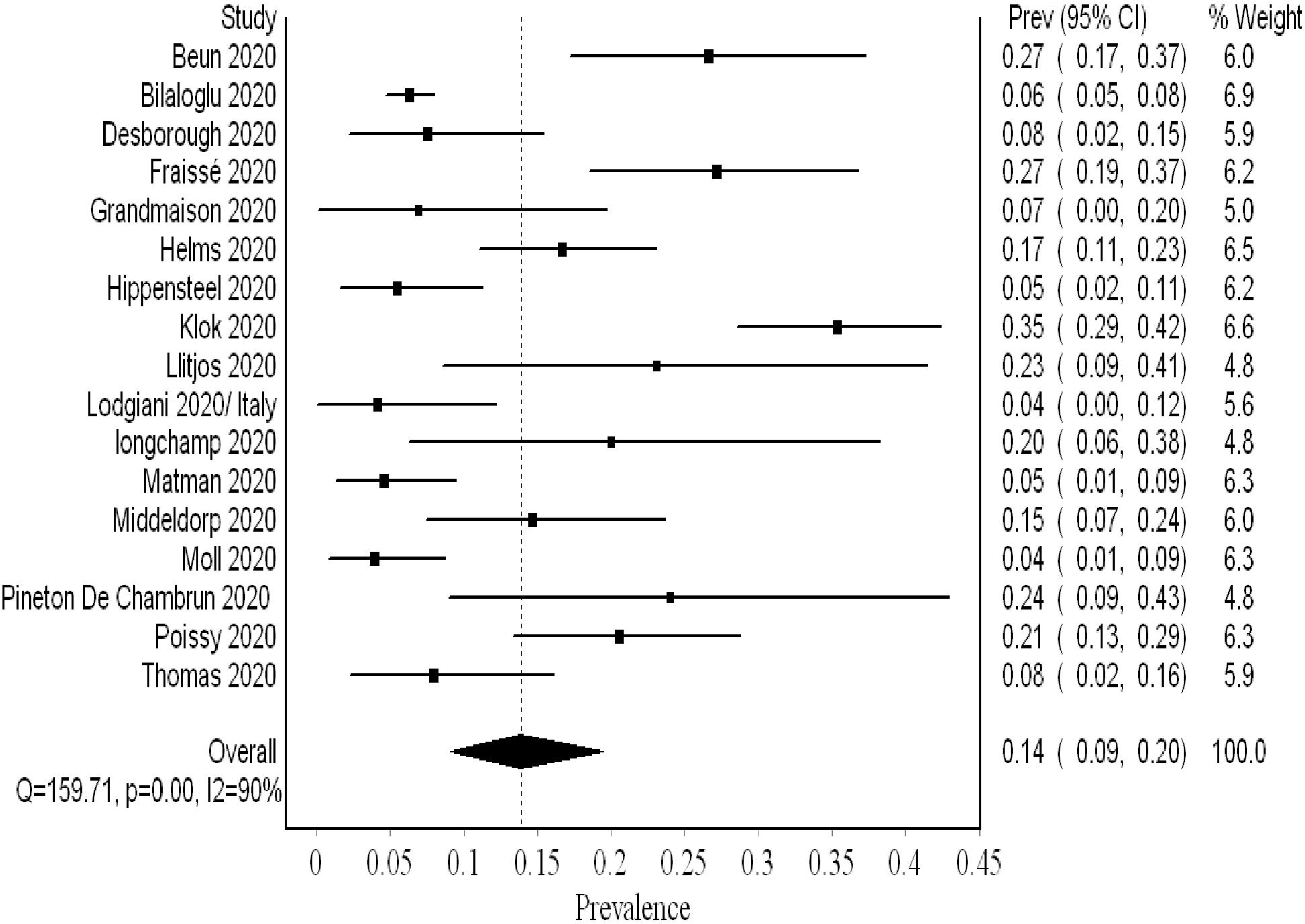
Forest plot showing the overall pooled proportion of PE

### Thromboprophylaxis strategy

Six studies reported the number of VTE events in patients receiving prophylactic anticoagulation (479 patients) compared to therapeutic dosages (83 patients). The dosages and definitions varied across these studies. In one study (pre- and post-intervention), a higher prophylactic dosage of nadroparin with adjustment guided by factor X-a activity (labeled as semi-therapeutic) was compared with standard prophylactic dose.^4,14,21^ For synthesis, we considered this adjusted dosage therapeutic and analyzed it in the corresponding arm (due to the paucity of studies). The VTE odds ratio was increased in the prophylactic anticoagulation group with uncertainty in the final point estimate OR 2.34 (95% CI 0.77, 7.14 I^2^ 53% Q 10). Three studies utilized systematic screening, hence, they provided a better estimate of the true VTE prevalence.^21^ We analyzed these studies separately, and the results showed significantly increased odds of VTE events with prophylactic dosing OR 5.45 (95% CI 1.90, 15.57 I^2^ 0% Q 1.2), and there was no evidence of heterogeneity (Figure 7).

**Figure 7:**
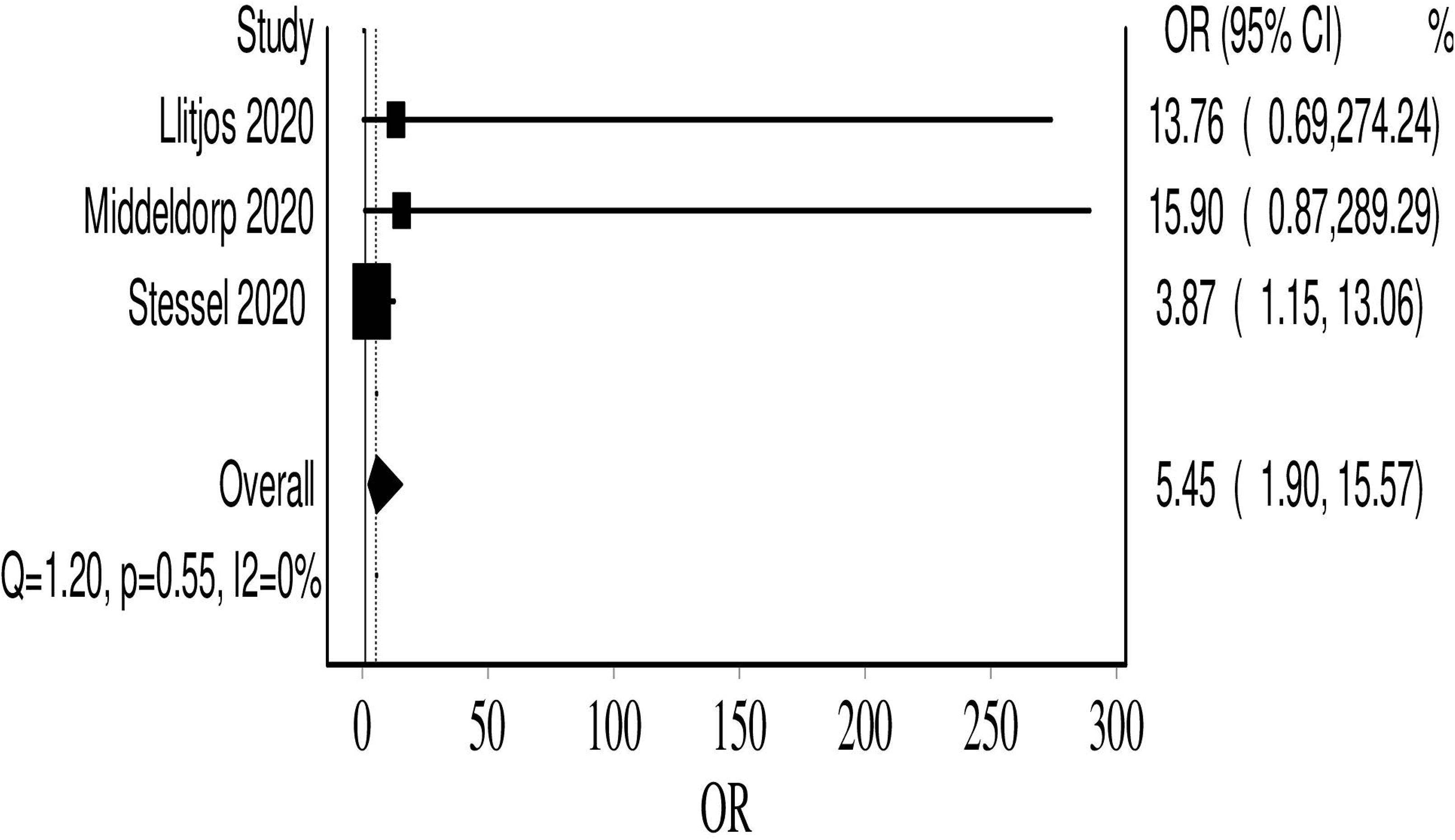
Forest plot showing the VTE event odds in the prophylactic anticoagulation group, compared to therapeutic dosing

### Quality assessment and risk of bias assessment

Most of the constituent studies had moderate or unclear risk of bias (Table 2). Although the number of included studies is adequate, the funnel plot suggested publication bias (Its value is limited in assessing prevalence studies publication bias). There was also reporting bias, as the reporting of distal DVT, PE, and VTE, method of diagnosis, dosing of chemoprophylaxis varied across studies.

**Table 2:** Table summarizing the risk of bias assessment

| Study | 1 | 2 | 3 | 4 | 5 | 6 | 7 | 8 | 9 |
| --- | --- | --- | --- | --- | --- | --- | --- | --- | --- |
| Al-samkari 2020 <sup>8</sup> | ? | + | + | - | - | - | - | - | - |
| Beun 2020 <sup>19</sup> | + | ? | + | - | - | + | - | - | - |
| Bilaloglu 2020 <sup>24</sup> | + | + | + | ? | - | + | - | - | - |
| Criel 2020 <sup>25</sup> | ? | ? | - | + | + | ? | + | + | - |
| Cui 2020 <sup>26</sup> | ? | ? | - | ? | + | ? | + | + | - |
| Desborough 2020 <sup>27</sup> | + | + | + | + | - | + | - | - | - |
| Fraissé 2020 <sup>28</sup> | + | ? | - | + | - | + | - | - | - |
| Grandmaison 2020 <sup>29</sup> | + | + | - | + | + | + | + | + | - |
| Helms 2020 <sup>30</sup> | + | + | - | + | - | + | - | - | - |
| Hippensteel 2020 <sup>9</sup> | + | + | - | + | - | + | - | - | - |
| Klok 2020 <sup>10</sup> | + | + | + | + | - | - | - | - | - |
| Llitjos 2020 <sup>4</sup> | + | + | - | + | + | + | + | + | - |
| Lodgiani 2020 <sup>11</sup> | + | + | + | + | - | + | - | - | - |
| longchamp 2020 <sup>12</sup> | ? | - | - | - | - | ? | + | - | - |
| Maatman 2020 <sup>13</sup> | + | + | - | + | - | + | - | - | - |
| Middeldorp 2020 <sup>14</sup> | + | + | + | + | + | + | + | - | - |
| Moll 2020 <sup>15</sup> | + | + | + | + | - | + | - | - | - |
| Nahum 2020 <sup>16</sup> | + | + | - | + | + | + | + | + | - |
| Pineton de Chambrun 2020 <sup>17</sup> | + | + | + | + | - | - | - | - | - |
| Poissy 2020 <sup>18</sup> | + | + | + | + | - | - | - | - | - |
| Ren 2020 <sup>20</sup> | + | + | + | + | + | + | + | + | - |
| Stessel 2020 1 <sup>21</sup> | + | + | + | + | ? | + | + | + | - |
| Stessel 2020 2 <sup>21</sup> | + | + | + | + | + | + | + | + | - |
| Thomas 2020 <sup>22</sup> | + | + | - | - | - | + | - | - | - |
| Zhang 2020 <sup>23</sup> | ? | + | + | - | ? | + | ? | + | - |

low risk;
high risk;
unclear risk assessment
Was the study's target population a close representation of the national population
ation in relation to relevant variables, e.g. age, sex, occupation?; (2) Was the sampling frame a true or close representation of the target population?; (3) Was some form of random selection used to select the sample, OR, was a census undertaken; (4) Was the likelihood of non-response bias minimal?; (5) Were data collected directly from the subjects (as opposed to a proxy); (6) Was an acceptable case definition used in the study?; (7) Was the study instrument that measured the parameter of interest (e.g. prevalence of low back pain) shown to have reliability and
validity (if necessary)?; (8) Was the same mode of data collection used for all subjects?; (9) Were the numerator(s) and denominator r(s) for the parameter of interest appropriate

## Discussion

Our meta-analysis comprised of over 2500 patients and revealed a high VTE prevalence of 0.31 (95% CI 0.24, 0.39) in critically ill COVID-19 patients. This prevalence increased to 0.48 (95% CI 0.33, 0.63) when systematic screening was utilized, meaning that almost one in two critical COVID-19 patients suffers from VTE. Furthermore, this heightened prevalence of VTE when systematic screening was used, did not include PE since it was not part of systematic screening. Hence, screening for PE systematically could have possibly further increased VTE prevalence. Even when nonsystematic screening was utilized, VTE prevalence remained high at 0.20 (95% CI 0.15, 0.26). Regarding PE and DVT prevalence, the overall prevalence of DVT (0.23) was higher than PE (0.14). This concurs with finding a high prevalence of undiagnosed DVT in autopsy evaluation of COVID-19 patients.^31^ Additionally, it may argue against the earlier literature suggesting that PE prevalence was much higher than DVT, proposing that PE events can originate in the lung’s vasculature in patients with SARS-CoV-2 infection.^32^

Our analysis revealed that approximately 40/100 additional DVTs are detected by systematic screening (0.48) compared to nonsystematic screening (0.08). This is likely due to the fact that asymptomatic DVT can be overlooked in nonsystematic screening. On the opposite side, PE is more likely to be associated with easily detected signs (sudden deterioration, unexplained tachycardia or sudden changes in the ventilator settings) especially in the context of ICU.

A recent study by Zhang et al. evaluated the utility of bedside ultrasonography in the diagnosis of DVT. It revealed a significantly higher DVT prevalence in deceased patients compared with surviving COVID-19 critically ill patients [94% (33/35) Vs. 47% (22/46), p < 0.001].^33^ Moreover, Wichmann et al. analyzed autopsies of 12 COVID-19 patients. They found that seven (58%) had undiagnosed VTE, while in four (33.3%), massive PE was the direct cause of death.^31^ Based on these data, we understand that the high mortality reported by many studies may actually be attributed to undiagnosed fatal VTE events. Consequently, studies with high mortality will likely underestimate the true VTE prevalence when deceased patients are excluded from screening. We additionally understand the impact of prevention and early identification on patient’s morbidity and mortality.

Tang et al. showed that prophylactic dosing of heparin in high-risk COVID-19 patients is associated with significantly lower mortality.^34^ This led the International Society on Thrombosis and Haemostasis (ISTH) amongst other societies to recommend a prophylactic dosage of pharmacological anticoagulants (LMWH or fondaparinux) for all hospitalized COVID-19 patients^3, 35^. However, it seemed that prophylactic anticoagulation is not sufficient for severe COVID-19 patients. This was concluded in a study by Llitjos and colleagues where they found a higher prevalence of VTE in patients on a prophylactic dose of anticoagulation (100%) compared to therapeutic anticoagulation (56%).^36^ More recently, Stessel et al. attempted the first quasi-experimental trial (pre- and post-intervention) comparing the mortality and incidence of VTE between conventional prophylaxis (once-daily nadroparin calcium 2850 IU) compared to an individualized semi therapeutic, prophylactic dosage guided by factor Xa activity (semi-therapeutic dosing). Both mortality (3.8% vs. 39.1%%, P<0.001) and VTE (15.3% vs. 41.3%, P=0.03) were significantly lower in the aggressive thromboprophylaxis group.^21^ Emerging evidence showed that even in COVID-19 patients receiving therapeutic anticoagulation, there is a high incidence of heparin resistance, and sub-optimal peak in anti-Xa levels.^37,38^ This may explain, in part, the high rate of VTE in patients on usual prophylactic doses and, even in patients on therapeutic dosing (although relatively at a lower rate).

Our review also aimed to address the uncertainty of using higher vs. standard prophylactic doses. When we limited our analysis to studies that only used systematic screening, and thus reduce the chances of missing fatal VTE events, we found that prophylactic dosing was associated with increased odds of VTE compared to therapeutic dosing (one study was counted in the therapeutic side although it used subtherapeutic dosing, due to limited studies).^21^ The results were homogenous. The reader should consider that the odds of VTE in the therapeutic arm were lower even in the likely event that those patients may have had VTE predisposing conditions, for which they were initiated on this therapeutic dosing (except Stessel et al.’s study, which was protocolized). This small unadjusted comparison suggests a value for a higher dosing or therapeutic chemoprophylaxis. Nonetheless, this will be confirmed by a number of ongoing trials aiming to address the efficacy and safety of various chemoprophylactic dosages (prophylactic, intermediates, weight-adjusted, or therapeutic); examples of such trials are IMPROVE (http://www.clinicaltrials.gov. NCT04367831), COVI-DOSE (http://www.clinicaltrials.gov. NCT04373707), and Hep-COVID (https://www.clinicaltrials.gov. NCT04401293). The safety of intensive thromboprophylaxis was not addressed in our review due to data paucity. Nonetheless, two recent observational studies suggested that this intensive thromboprophylaxis is safe in terms of inducing major bleeding events. ^39,40^ Thus, we believe that the intensive thromboprophylaxis protocol suggested by Stessel et al. is a valid chemoprophylaxis regimen until further data from ongoing RCTs becomes available.^21^

Limitations of our review are the heterogeneity in the pooled prevalence in the constituent studies. This is likely due to varying detection methods (systematic vs. nonsystematic, imaging modalities used, timing, etc.), screening threshold (many studies reported that the threshold was high due to infection control concerns), varying severity of illness, prophylaxis strategies, and dosage, missing VTE in deceased patients of fatal VTE events, varying and insufficient follow-ups. Additionally, the inability to provide a mortality comparison between VTE group and non-VTE group due to data paucity (we contacted the primary authors; however, we could not get the data necessary for its computation), limited conclusion provided by the comparison of VTE in the therapeutic vs. prophylactic anticoagulation groups (absence of adjustment, varying doses between studies). Moreover, the retrospective nature of the included studies, inability to accurately compute the prevalence of PE (absence of systematic PE screening), and absence of autopsies to ascertain causes of death, adds to the limitations of our review.

Notwithstanding this, there are many strengths to our review that are worthy of mention. This is the most extensive review examining the prevalence of VTE exclusively in critically ill patients. Additionally, the review examines VTE prevalence based on the utilized screening method providing the readers with a better estimate of VTE prevalence. We also pooled a proportion that reflects the prevalence; nonetheless, we acknowledged its limited accuracy. Finally, we believe that the results of the limited comparison between lower and higher dosing of chemoprophylaxis will help inform therapeutic decisions until further data from RCTs becomes available.

Future research direction should evaluate the utility of systematic screening and early therapeutic anticoagulation dosage on outcomes (VTE progression, ICU stay, and mortality). The utility of systematic screening with US at regular intervals to ascertain the exact prevalence of VTE is needed. In these studies, patients with distal DVT should be temporally followed and compared to a non-DVT cohort, to determine the incidence of proximal DVT, PE, and mortality events. This will ascertain the exact need for therapy in these patients.

In conclusion, our review of critically ill COVID-19 patients revealed a high prevalence of VTE events. This prevalence is higher when systematic screening is utilized. Our review concluded a potential for higher prophylactic or therapeutic dosages in reducing VTE burden. Data from ongoing RCTs are awaited to further confirm the findings of our review.

## Data Availability

Data are available upon request from the corresponding author.

## Declarations

### Availability of data

Data are available upon request from the corresponding author.

### Author’s contribution

MFHM conceived the idea of the review and formed the team. MFHM conducted the initial search, and with SFM conducted the screening. MFHM, KMS, SDA, SFM, SII, MSN and LOA extracted the data. MFHM performed the analysis. MFHM constructed the tables and figures and wrote the initial draft; the manuscript was then critically reviewed and revised by all the study 2authors. The final version was approved by all authors for publication.

### Ethics approval

Formal ethical approval was not needed for our review since it is a synthesis of already available data.

### Consent to participate

Not applicable

### Consent for publication

All authors approved the final version of the review for publication

## Acknowledgment

We acknowledge the editor and the reviewers for the timely review and constructive feedback.

